# Anemia, hemoglobin concentration and cognitive function in the Longitudinal Ageing Study in India-Harmonized Diagnostic Assessment of Dementia (LASI-DAD) and the Health and Retirement Study

**DOI:** 10.1101/2024.01.22.24301583

**Authors:** Laura M. Winchester, Danielle Newby, Upamanyu Ghose, Peifeng Hu, Hunter Green, Sandy Chien, Janice Ranson, Jessica Faul, David Llewellyn, Jinkook Lee, Sarah Bauermeister, Alejo Nevado-Holgado

## Abstract

**Background:** In India, anemia is widely researched in children and women of reproductive age, however, studies in older populations are lacking. Given the adverse effect of anemia on cognitive function and dementia this older population group warrants further study. The Longitudinal Ageing Study in India – Harmonized Diagnostic Assessment of Dementia (LASI-DAD) dataset contains detailed measures to allow a better understanding of anaemia as a potential risk factor for dementia.

**Method:** 2,758 respondents from the LASI-DAD cohort, aged 60 or older, had a complete blood count measured from venous blood as well as cognitive function tests including episodic memory, executive function and verbal fluency. Linear regression was used to test the associations between blood measures (including anemia and hemoglobin concentration (g/dL)) with 11 cognitive domains. All models were adjusted for age and gender with the full model containing adjustments for rural location, years of education, smoking, region, BMI and population weights.

Results from LASI-DAD were validated using the USA-based Health and Retirement Study (HRS) cohort (n=5720) to replicate associations between blood cell measures and global cognition.

**Results:** In LASI-DAD, we showed an association between anemia and poor memory (p=0.0054). We found a positive association between hemoglobin concentration and ten cognitive domains tested (β=0.041-0.071, p<0.05). The strongest association with hemoglobin was identified for memory-based tests (immediate episodic, delayed episodic and broad domain memory, β=0.061-0.071, p<0.005). Positive associations were also shown between the general cognitive score and the other red blood count tests including mean corpuscular hemoglobin concentration (MCHC, β=0.06, p=0.0001) and red cell distribution width (RDW, β =-0.11, p<0.0001). In the HRS cohort, positive associations were replicated between general cognitive score and other blood count tests (Red Blood Cell, MCHC and RDW, p<0.05).

**Conclusion:** We have established in a large South Asian population that low hemoglobin and anaemia are associated with low cognitive function, therefore indicating that anaemia could be an important modifiable risk factor. We have validated this result in an external cohort demonstrating both the variability of this risk factor cross-nationally and its generalizable association with cognitive outcomes.

## Introduction

Anemia has been associated with poor cognition and physical performance ^1^ and is characterised by low hemoglobin concentration. Low hemoglobin is associated with poor cognition and dementia in western elderly populations ^2^. In 2021, the total prevalence of anemia in South East Asia was 43% in contrast to western regions, such as 6.8% in North America ^3^. Although, anemia is widely researched in childhood development and among women of reproductive age in India ^4^, studies in older populations are lacking. A better understanding of anemia and the effects of hemoglobin concentration on cognitive function will allow clinicians and researchers to make informed recommendations on suitable interventions such as dietary control or supplements in this high-risk population. Examination of the complete blood panel enables us to study underlying conditions and get a broader view on changes in the red blood indices and their relationship to cognition.

Deficits in cognition serve as a potential indicator of dementia and have been used in research as a proxy for early changes prior to clinical symptoms. Dementia prevention is increasingly prioritised as an approach to reduce the impact of dementia globally, particularly in low and middle income countries ^5^.

The Longitudinal Ageing Study in India – Harmonized Diagnostic Assessment of Dementia (LASI-DAD) is an observational cohort collected as part of a larger longitudinal ageing study (LASI) ^6^. LASI-DAD contains more detailed phenotyping including a battery of cognitive function tests, comprehensive geriatric assessments and venous blood collection. This blood collection from over 2892 respondents enables a range of clinical tests, genetic assays, biomarkers and complete blood count to be measured. These detailed measures make the LASI-DAD an ideal cohort to better understand the potential impact of anemia and changes in blood hemoglobin and cognition in an elderly dataset from India.

In this study, we aimed to determine the relationship between anemia and cognitive function. The objectives of this study were firstly to characterise population to understand the distribution of hemoglobin and red blood cell related measures and secondly to determine the relationship between hemoglobin and related blood measures with cognitive function. Finally, we validated the results from LASI-DAD using an external cohort from the USA based Health and retirement study with a similar age distribution. These datasets will allow us to look for markers of change that are not related to cultural differences and present traits unique to the Indian population.

## Methods

### Longitudinal Ageing Study in India – Harmonized Diagnostic Assessment of Dementia (LASI-DAD) Study population

#### Respondents

The Longitudinal Ageing Study in India – Harmonized Diagnostic Assessment of Dementia (LASI-DAD) is a sub-study of the larger LASI study. The LASI study itself is the largest of the Health and Retirement Family of cohorts with 72,000 respondents, gathering information at household and individual levels and representative of both the states and national population^7^. The LASI-DAD subset of respondents includes detailed demographics and lifestyle interviews as well as detailed cognitive function testing (n = 4096). LASI-DAD is observational cohort study with data collection ongoing ^8^. This study focuses on the Wave 1 baseline population with venous blood samples which was collected from 2017 and 2020 (LASI-DAD version A.3). Respondents were aged 60 or older and residents of 18 states and union territories across India. From the initial LASI-DAD population, all were eligible for the blood assays, but only 2892 consented and had venous blood samples taken. From these samples, a biomarker panel and complete blood count was measured ^9^. Samples were excluded from this analysis if they had self-reported diseases related to blood loss (dengue fever or chikungunya) in the last 2 years or were admitted to the hospital in the last 12 months with vector-born disease (Chikungunya, Filariasis, Dengue), liver diseases (hepatitis, alcoholic liver disease, cirrhosis) or malaria resulting in a final population of 2854 for analysis.

#### Variables

##### Hemoglobin and Complete blood count measures

Complete blood count measures were analysed from 2,892 venous blood samples assayed at the Metropolis pathology laboratory on the Beckman Coulter LH780. This standard panel of red and white blood cell measures included red blood cell count (RBC), hemoglobin concentration (HGB), and derived parameters such as red cell distribution width (RDW) and mean corpuscular hemoglobin concentration (MCHC). Inclusion of the full blood count panel allowed the assessment of anaemia subtypes and derived hemoglobin variables as potential proxies for iron markers.

Hemoglobin concentration was used to define anemia as specified by World Health Organisation (WHO) diagnostic guidelines, whereby for males a concentration of < 13 g/dL was considered anaemic and in females < 12 g/dL ^10^. Additional subgroups of severity were defined as moderate anemia (13g/dL / 12g/dL - 11g/dL), moderate - severe anemia (8g/dL - 11g/dL) and severe anemia (< 8g/dL) according to Northrop-Clewes *et al.* ^11^. Microcytic anemia and Macrocytic anemia were defined as MCV < 83fL and MCV > 101fL respectively.

##### Cognitive tests

Eleven summary scores were used to represent different cognitive phenotypes. Tests were organised into broad domains which included orientation, executive functioning, language/fluency, memory, and visuospatial to assess respondent ability as described by Lee at al. ^12^. This cognitive test battery is designed to assess mild cognitive impairment and dementia and is based on the Harmonized Cognitive Assessment Protocol (HCAP). The HCAP battery was modified as needed to be culturally sensitive and appropriate for local contexts and population characteristics and included additional tests suitable for illiterate/innumerate respondents. Assessors administered the domain-specific tests, and interviewed a family member or friend, whom study respondents nominated as informants. The main summary score is a general cognitive factor score consolidating memory, executive functioning, visuospatial, and language domains ^13^. Informant reporting was used define the respondent group who were diagnosed with memory problems.

##### Covariates

In all models, age at baseline and sex are included as covariates. Years of education is included as a confounder for the cognition outcome. Other demographics included as confounders were location (rural or city), region (defined by orientations of location by pole e.g. north/south, Supplementary Table 1), Body Mass Index (BMI) and smoking status (non-smokers, previous smokers and current smokers).

### Health and Retirement Study

#### Respondents

The Health and Retirement Study (HRS) was used to compare and validate blood count associations found in this paper focussing on LASI-DAD. The HRS is nationally representative of the USA and has been collecting respondent data on economic and health since 1992 ^14^. A cross-sectional subset of HRS (RAND HRS is 2020v1. Harmonized HRS is D); Wave 13 (2016) of data collection was used as this incorporates the biomarker panel, cognitive tests and demographics necessary for replication of our study in LASI-DAD. Respondents under 60 years old were excluded resulting in 7598 samples for analysis.

#### Variables

##### Global cognition

A total cognitive score, as well as a total word recall summary and total mental status summary scores (n = 7598) were used to assess cognitive function in the respondents. The total cognitive score is comprised of a total word recall and mental status summary scores (range = 0-35). Total word recall summary is comprised of immediate and delayed word recall tests. The mental health score combines the following tests: serial 7’s, object, date, backwards counting from 20, and President/Vice-President naming tasks. Imputed scores were used from each ^15^ to reduce the bias from missingness due to item non-response, as cognitively impaired respondents are less likely to complete the cognitive assessment (Rogers et al., 2009).

##### Red blood cell measures

Wave 13 of the HRS cohort contains biomarker assays from venous blood samples of respondents. This contains the equivalent panel of blood cell measures as measured in the LASI-DAD cohort. Assays were carried out at the University of MN Advanced Research and Diagnostic Laboratory (ARDL) on a Sysmex XE-2100 instrument.

##### Covariates

As described with the LASI-DAD cohort all models were corrected for age and sex. The other confounders considered were census region, BMI, smoking, ethnicity and years of education.

### Statistical Methods

All analysis was performed using R version 4.2.2. Descriptive statistics were calculated to characterize the study cohort with respect to all study variables. Differences between sex were assessed by t-test and chi-squared tests as appropriate. The LASI-DAD population was split into age subgroups by 10 year age bands.

#### Regression Models

For the main analysis, general linear regression models were used to estimate the association between each blood measure (exposure) and cognitive function test (outcomes) while adjusting for confounders. Logarithm transformations were applied to standardise continuous variables for effect size interpretation. We used three models to understand how population specific confounders might impact the relationship between hemoglobin and cognition. Model 1 was adjusted for sex and age only whereas Model 2 additionally including years of education, rural area, BMI, geographic region, and smoking. Included in Model 3 was a sampling weight to allow for the difference between selection strategies and non-responders between the main population of the same age and the biomarker assessed samples. Sampling weights were calculated using sex, marital status, education, parent’s education, literacy status, binary indicators for state of residence, rural area, caste, household income, wealth quintiles and health variables ^12^. All analyses were performed on the whole sample and stratified by sex. P-values were adjusted for multiple testing using Benjamini and Hochberg.

## Results

### Anemia in the LASI-DAD Study population

2,898 of the LASI-DAD respondents had complete blood count results. From 2,758 LASI-DAD respondents who were eligible for analysis, 52.6% were female. These respondents had a mean age of 69.5 years and mean of 3.83 years of education. 39% (n = 1112) of the measured population were anaemic according to WHO diagnostic guidelines. Hemoglobin concentration varied by sex (p-value < 1 x 10^-16^), with males having higher hemoglobin levels than females (Table 1). Large sex-based differences were seen for age, years of education, BMI and smoking (p-value < 1.01 x −10-14). There were smaller differences in region and urban location by sex (p-value = 0.041 and 0.038).

**Table 1:**
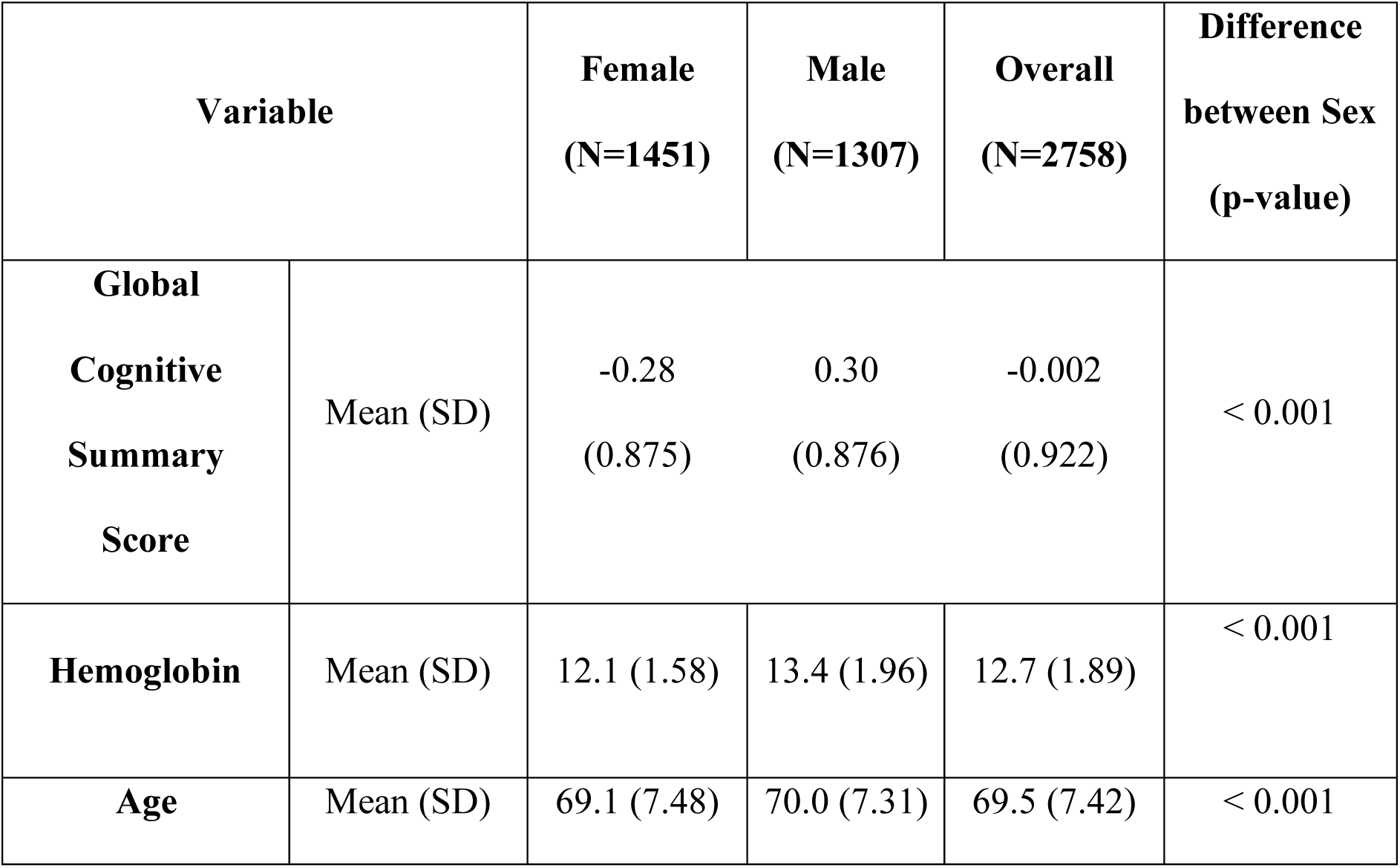
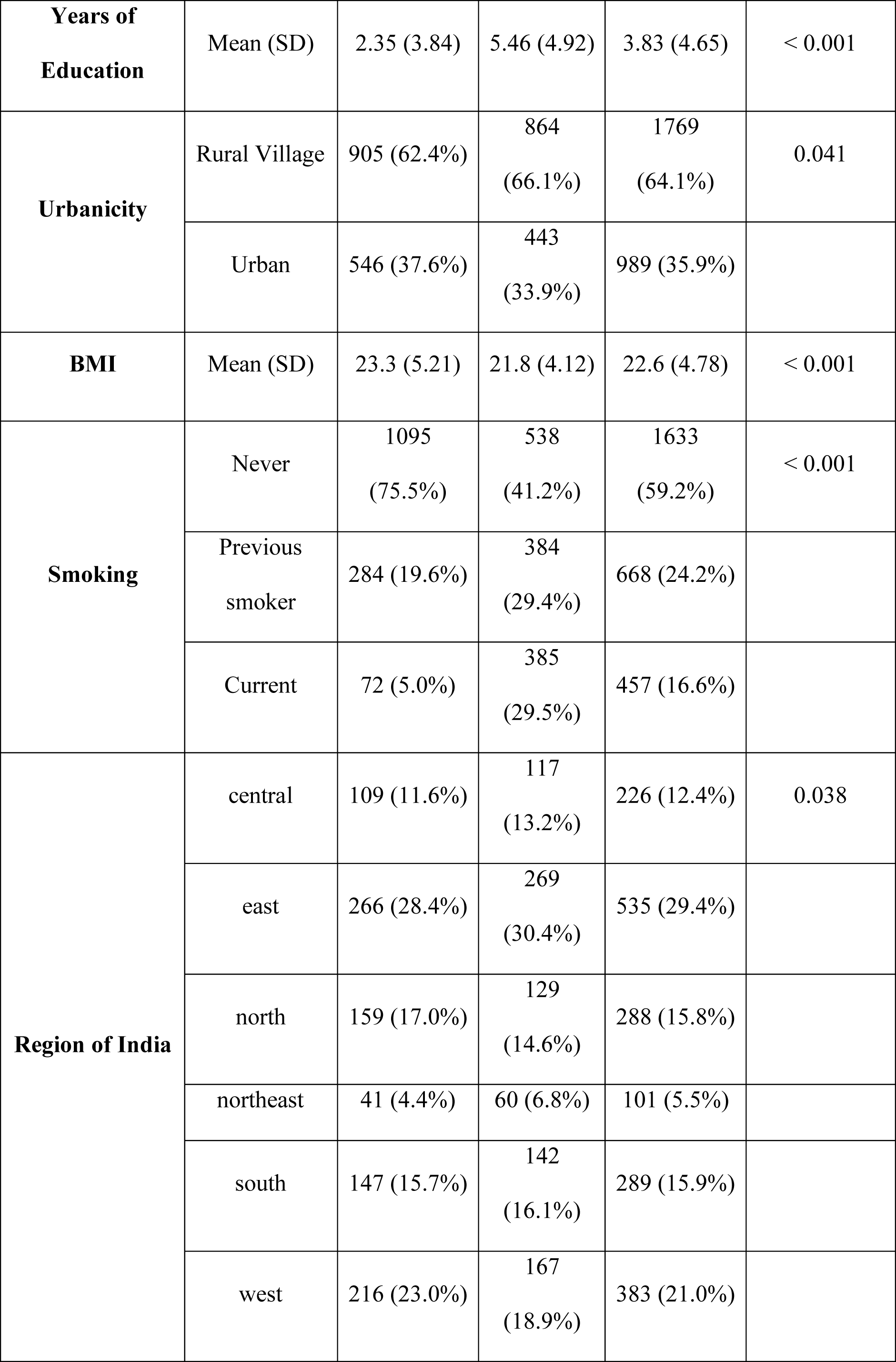
Demographics of the LASI-DAD Venous Blood sample sub-cohort.

| Variable |  | Female<br>(N=1451) | Male<br>(N=1307) | Overall<br>(N=2758) | Difference<br>between Sex<br>(p-value) |
| --- | --- | --- | --- | --- | --- |
| **Global Cognitive Summary Score** | Mean (SD) | -0.28<br>(0.875) | 0.30<br>(0.876) | -0.002<br>(0.922) | < 0.001 |
| **Hemoglobin** | Mean (SD) | 12.1 (1.58) | 13.4 (1.96) | 12.7 (1.89) | < 0.001 |
| **Age** | Mean (SD) | 69.1 (7.48) | 70.0 (7.31) | 69.5 (7.42) | < 0.001 |
| **Years of Education** | Mean (SD) | 2.35 (3.84) | 5.46 (4.92) | 3.83 (4.65) | < 0.001 |
| **Urbanicity** | Rural Village | 905 (62.4%) | 864<br>(66.1%) | 1769<br>(64.1%) | 0.041 |
|  | Urban | 546 (37.6%) | 443<br>(33.9%) | 989 (35.9%) |  |
| **BMI** | Mean (SD) | 23.3 (5.21) | 21.8 (4.12) | 22.6 (4.78) | < 0.001 |
| **Smoking** | Never | 1095<br>(75.5%) | 538<br>(41.2%) | 1633<br>(59.2%) | < 0.001 |
|  | Previous smoker | 284 (19.6%) | 384<br>(29.4%) | 668 (24.2%) |  |
|  | Current | 72 (5.0%) | 385<br>(29.5%) | 457 (16.6%) |  |
| **Region of India** | central | 109 (11.6%) | 117<br>(13.2%) | 226 (12.4%) | 0.038 |
|  | east | 266 (28.4%) | 269<br>(30.4%) | 535 (29.4%) |  |
|  | north | 159 (17.0%) | 129<br>(14.6%) | 288 (15.8%) |  |
|  | northeast | 41 (4.4%) | 60 (6.8%) | 101 (5.5%) |  |
|  | south | 147 (15.7%) | 142<br>(16.1%) | 289 (15.9%) |  |
|  | west | 216 (23.0%) | 167<br>(18.9%) | 383 (21.0%) |  |

There was a significant difference between age and hemoglobin (p-value < 1 x 10^-16^). To better understand this association, we split the sample into 10 year groups and compared hemoglobin and its derived clinical phenotypes. Standard anemia is more common in older respondents in this population (Table 2). Anemia severity subtypes derived from complete blood count measures showed the same proportional increase. In contrast, there was no significant difference in the proportion of microcytic anaemia respondents (p>0.05). Macrocytic individuals were rarer in the population (0.1%)

**Table 2:** Distribution of the hemoglobin concentration and related conditions by age. Age is split into 10 year bins to understand the changes in hemoglobin during ageing in the LASI-DAD cohort. Subtypes were defined as the following: Moderate anemia = 13g/dL / 12g/dL - 11g/dL; Moderate - severe anemia = 8g/dL - 11g/dL; Severe anemia < 8g/dL; Microcytic anemia = MCV < 83fL Macrocytic anemia = MCV > 101fL

|  | **60 - 70** | **70 - 80** | **80 - 90** | **90 + years** | **Overall** |
| --- | --- | --- | --- | --- | --- |
|  | **years** | **years** | **years** | **(N=48)** | **(N=2758)** |
|  | **(N=1600)** | **(N=843)** | **(N=267)** |  |  |
| **Hemoglobin Concentration** |  |  |  |  |  |
| Mean (SD) | 12.8 (1.85) | 12.6 (1.95) | 12.3 (1.97) | 11.6 (1.89) | 12.7 (1.91) |
| Median [Min, Max] | 12.9 [3.80, 20.5] | 12.6 [4.60, 18.8] | 12.6 [4.40, 17.2] | 11.9 [4.70, 15.2] | 12.7 [3.80, 20.5] |
| **Anemia** | 581<br>(35.0%) | 370<br>(42.6%) | 130<br>(47.1%) | 31 (60.8%) | 1112<br>(39.0%) |
| **Moderate Anemia** | 352<br>(21.2%) | 214<br>(24.7%) | 74 (26.8%) | 14 (27.5%) | 654<br>(22.9%) |
| **Moderate to Severe Anemia** | 182<br>(11.0%) | 129<br>(14.9%) | 41 (14.9%) | 12 (23.5%) | 364<br>(12.8%) |
| **Severe Anemia** | 24 (1.4%) | 16 (1.8%) | 11 (4.0%) | 3 (5.9%) | 54 (1.9%) |
| **Macrocytic Anemia** | 1 (0.1%) | 2 (0.2%) | 0 (0%) | 0 (0%) | 3 (0.1%) |
| **Microcytic Anemia** | 375<br>(22.6%) | 168<br>(19.4%) | 52 (18.8%) | 13 (25.5%) | 608<br>(21.3%) |

### Anemia is associated to lower cognitive domain scores

Respondents classified as anaemic were associated with the group diagnosed with memory conditions (p-value = 0.0054). We found a larger effect on the memory score compared to general cognition score for all anemia severity subgroups (Supplementary Figure 2). Moderate to severe anemia had the strongest association with memory score (beta = −0.18, p-value > 0.05).

### Association between hemoglobin concentration and harmonised cognitive test measures

Given the reported relationship between anaemia (as a clinical classification) and cognition we wanted to better understand the phenotypes by examining haemoglobin as a continuous measure. There was a significant association between lower hemoglobin levels and poorer performance on cognitive tests (Figure 1). The strongest associations were identified for the immediate and delayed episodic memory-based subdomain measures. This was supported by the other cognitive domains tested whereby ten out of the eleven measures tested were statistically significant apart from the narrow domain recognition memory. Three models were tested to understand how confounders might impact the key association between hemoglobin and global cognition (Table 3). When the model was corrected for known population confounders (education, region, and rural location) as well as exposure confounders (BMI, smoking) the associations remained (Model 2), although beta effects were attenuated with the increased number of covariates. Associations also remained after adjustment for sample weights (Model 3).

**Figure 1.**
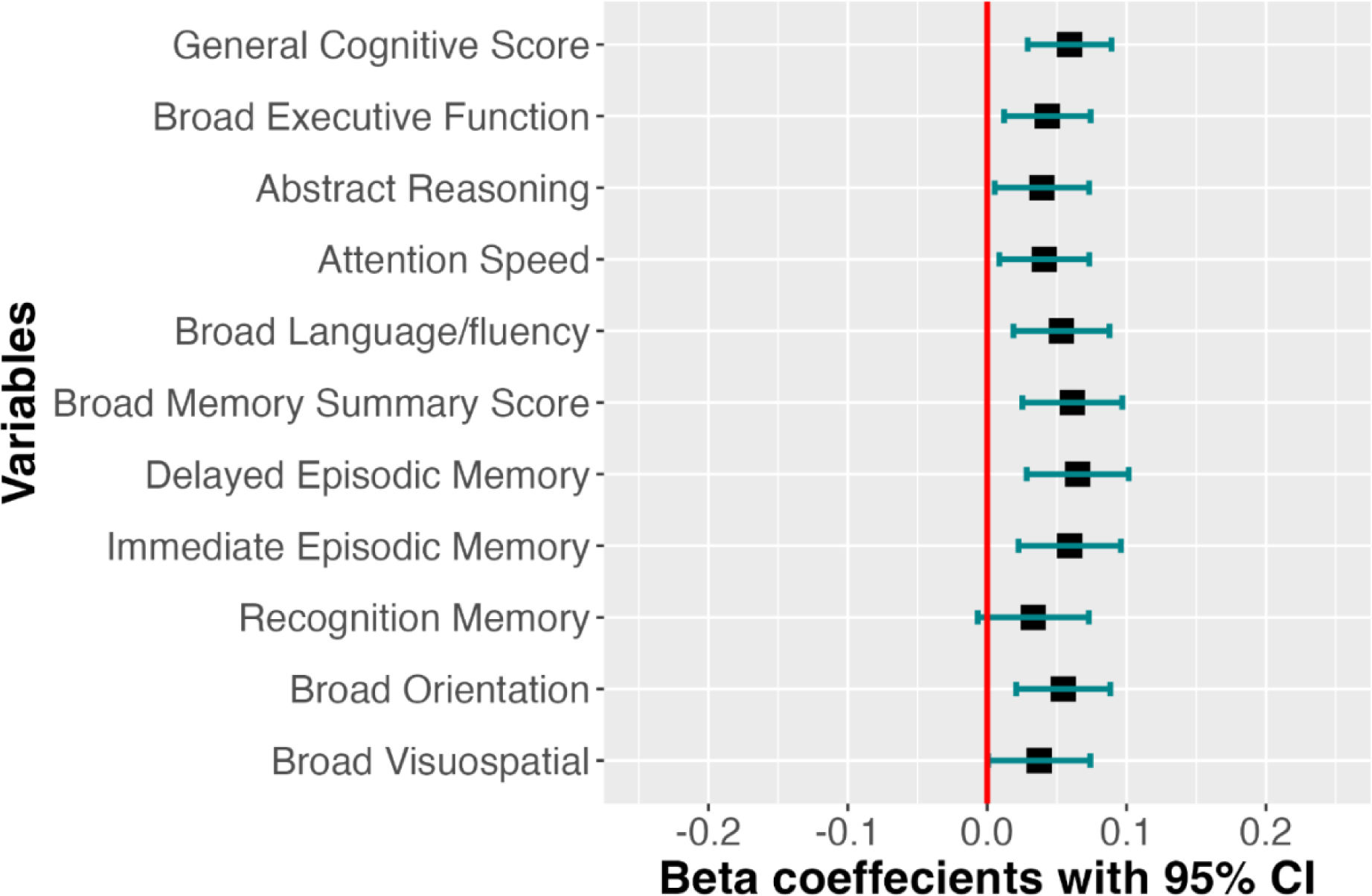
Association between hemoglobin and general cognition in the LASI cohort. Results from regression Model 2 for cognitive domains (hemoglobin + age + gender + rural location + region + BMI + smoking + ethnicity + years of education). Ten cognitive tests described in the LASI-DAD cohort had a significant association to hemoglobin.

**Table 3.** Comparison of Models showing the association between hemoglobin and global cognition.

| **Model** | **Standardised**<br>**beta** | **Lower**<br>**95% CI** | **Upper**<br>**95% CI** | **P value** |
| --- | --- | --- | --- | --- |
| Model 1 | 0.10 | 0.06 | 0.13 | < 0.001 |
| Model 2 | 0.06 | 0.04 | 0.09 | < 0.001 |
| Model 3 (including sample weighting) | 0.07 | 0.04 | 0.10 | < 0.001 |

Due to the difference in classification of anemia between sexes, we repeated the analysis using a dichotomous indicator and stratifying by sex. Anemia was associated with poor cognitive function in both men and women, with a common direction and overlapping confidence interval regions (Table 4).

**Table 4.** Sex based Stratification of Model 1 showing the association of global cognition to hemoglobin.

|  | **Male** |  |  | **Female** |  |  |
| --- | --- | --- | --- | --- | --- | --- |
|  | **Beta** | **CI** | **p-value** | **Beta** | **CI** | **p-value** |
| Hemoglobin | 0.12 | 0.06 – 0.17 | <0.001 | 0.08 | 0.03 – 0.13 | 0.002 |
| Age | -0.24 | -0.29 – -0.18 | <0.001 | -0.31 | -0.36 – -0.26 | <0.001 |

### Relationship between cognitive function and other red blood count measures

There were significant associations found between the general cognitive score and the other red blood count tests (Figure 2). RBC, hemoglobin and MCHC all showed a significant positive associations with the general cognitive score. RDW was most strongly associated with global cognition (β = −0.07, p < 0.0001).

**Figure 2:**
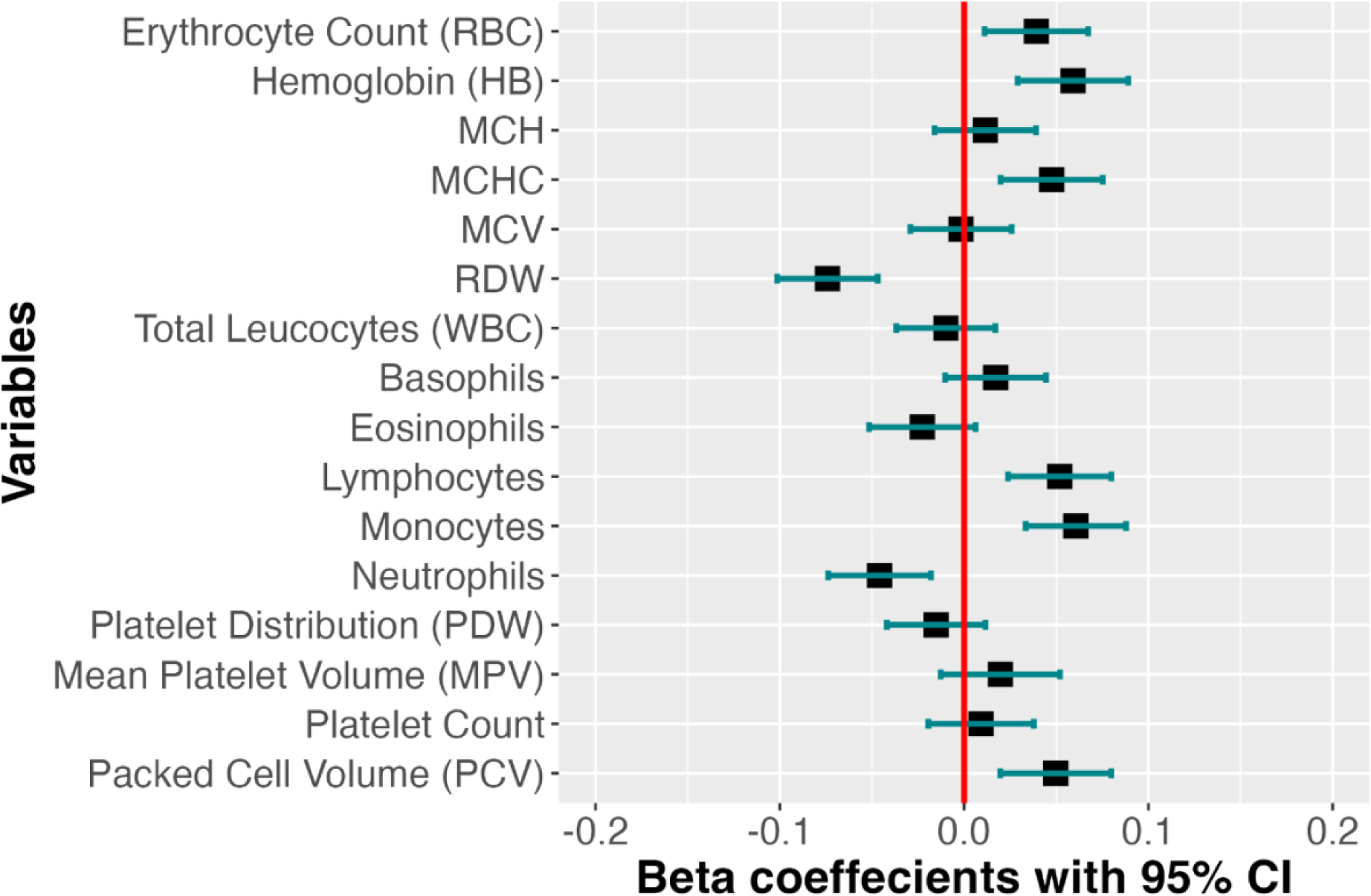
Association between total cognition and red blood indices in the LASI cohort. Results from regression Model 2 for general cognitive score (blood measure + age + gender + rural location + region + BMI + smoking + ethnicity + years of education). Seven out of the 16 blood indices had an association to global cognition in the sub-cohort. This included white and red measures although strongest effects were shown in the red blood phenotypes (RDW and HGB). RDW = red blood cell distribution width, MCHC = mean corpuscular hemoglobin concentration

We also found significant negative associations between the general cognitive score with three of the white blood cell measures (Figure 2), including monocytes and lymphocytes (β = 0.04, p < 0.05) and neutrophils (β = −0.03, p = 0.0212).

### Validation of the association between hemoglobin and cognitive function in the Health and Retirement Study

Respondents selected from the HRS were aged between 61 and 107 (mean = 72.7 years) and located across the USA (Supplementary Table 2). 57.7% of the respondents were female with a mean of 12.8 years of education. The proportion of respondents with anemia was less than the LASI cohort, with only 20.3% with a clinical diagnosis of anemia (39% in LASI-DAD, Table 1). Severe anemia was also less prevalent in the HRS wave 13 (0.2% in HRS, 1.9% in LASI-DAD).

In HRS, global cognition score showed a significant association between lower cognition and hemoglobin concentration (Supplementary Table 3). Using the fully adjusted model (Model 2), the total cognitive summary score showed a significant association with all red blood count measures (Figure 3). Here, all hemoglobin derived measures were associated with cognitive outcomes with hemoglobin concentration having the strongest effect on global cognition (β = 0.077 p < 10^-8^). As with LASI-DAD, a negative relationship was found between RDW and cognition (β = 0.077 p < 10^-8^). White blood cell measures reflected the associations seen in LASI-DAD with significant associations detected between general cognitive score and lymphocytes, monocytes and neutrophils (β = 0.043, 0.029, −0.043, p < 0.001).

**Figure 3.**
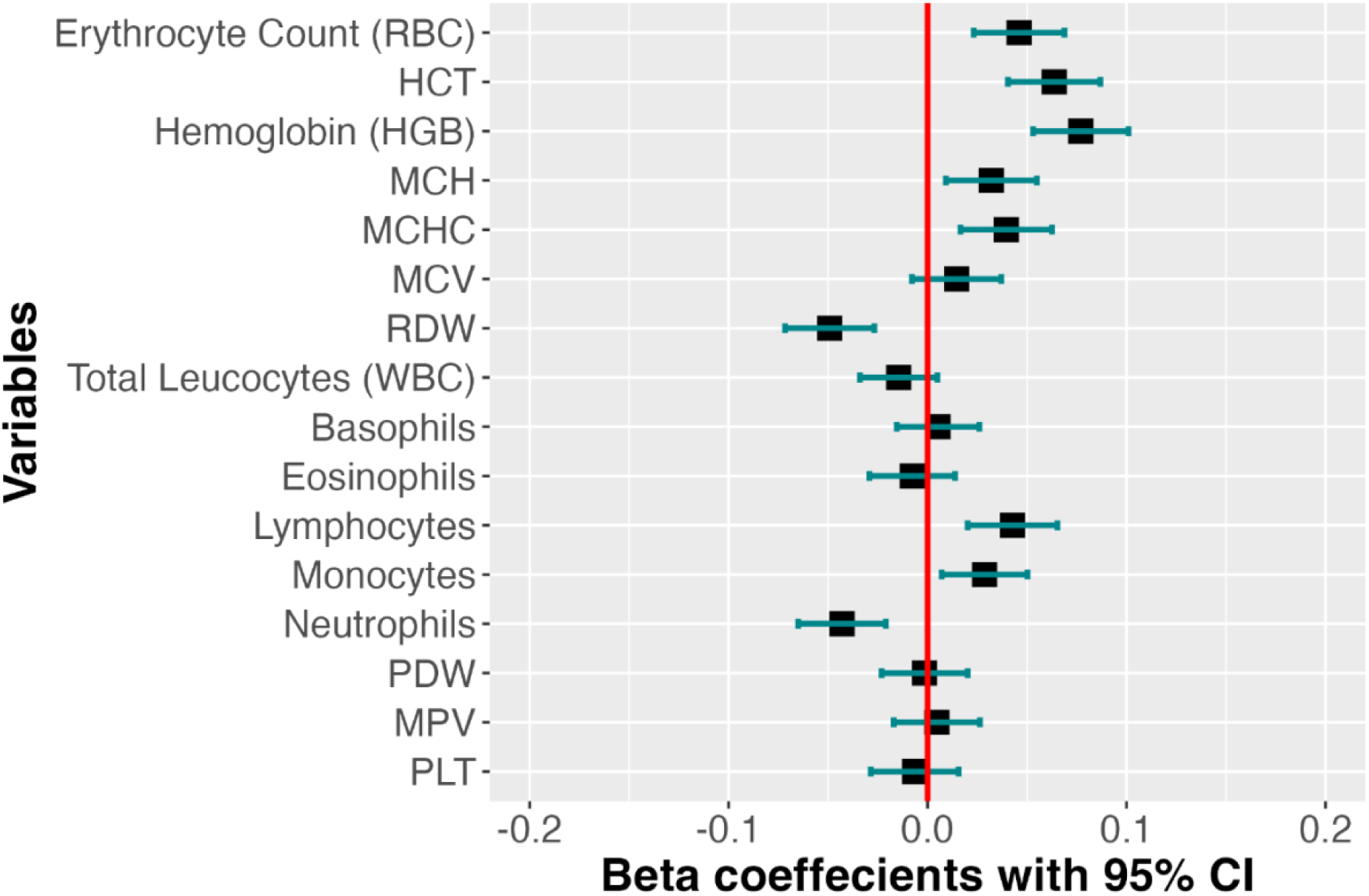
Blood indices were also associated in cognition in the Health and Retirement study validation cohort. Results from regression Model 2 for global cognitive score (blood measure + age + gender + census region+ BMI + smoking + ethnicity + years of education). Blood indices measured were RBC = red blood cell, RDW = red blood cell distribution width, MCHC = mean corpuscular hemoglobin concentration, MCH = mean corpuscular hemoglobin, HGB = hemoglobin concentration, HCT = hemocrit, PLT = Platelet Count, MPV = Mean Platelet Volume, PDW = Platelet Density Width

## Discussion

This study describes the distribution of anemia and cognition in a representative sample of older Indians. The percentage of study respondents (38%) with low hemoglobin (anemia) is lower compared to the prevalence for the South Asian region (43%). However, as reported elsewhere, we found females had an increased prevalence of anemia compared to males ^16^. We showed the association between low cognition functioning across a variety of domains and low hemoglobin concentration. After correction for confounding factors and validation in an external cohort, the association was still statistically significant indicating the robustness of the results. This association is also supported in the associated red blood count. Although the inclusion of some of variables has reduced the effect on the outcome it was important to assess the impact of both region and rural location on the results due to underlying confounders described in the cohort previously ^17^. The main association between hemoglobin concentration and global cognition was robust to model variations and adjustment for additional covariates and possible confounders.

Using the data on complete blood count we detected associations between multiple blood count measures and cognitive function. We detected a strong association between RDW and cognition in both LASI-DAD and HRS. RDW is used clinically to help define anemia subtypes, although it may also be predictive of cardiovascular and cancer related diseases. This measure has been associated with dementia in an East Asian cohort from China ^18^. RDW has been associated with ageing, more generally, in studies using genetics to better understand the mechanism of the age-related health changes ^19^. Higher white blood indices are associated with an inflammatory response. For example, the association between cognition and neutrophils, found in both LASI-DAD and HRS, may indicate activation of inflammatory processes and Alzheimer’s Disease progression ^20^.

Previous studies have reported associations between anaemia and particular cognitive domains^21^. We also see a significant association between executive function domains and memory. The tests representing memory scores for episodic memory showed the largest adverse effect from reduced hemoglobin. This study uses measures of cognitive scores as a proxy for cognitive change. Cognitive change in a cohort of this age group is likely to linked to Mild Cognitive Impairment and therefore potential dementia cases ^22^.

Implicit to study design was the use of multiple cohorts to allow us to understand more about population specific effects on anemia and red blood cell measures. Association between anemia and cognition was demonstrated between both populations. Given the increased prevalence of anemia in the LASI-DAD cohort it is important to understand the impact of the changes on cognition. However, the commonality of directional effect between the cohorts analysed means that the changes measured are ideal as a cross population marker. Here, we described two distinct cohorts with common cognitive test domains. We used general cognitive score for comparing to blood count measures and across cohorts as this had a high marginal reliability between cohorts when harmonised ^23^. Additionally, a cross sectional study of the complete blood count in the UK Biobank cohort showed similar associations between cognition and complete blood count measures ^24^.

### Strengths and Limitations

This study uses two independent cohorts from the HRS family which allows us to compare both hemoglobin exposures and cognitive outcomes to generate a deeper picture of the relationship between the two measures. Although datasets were not fully harmonised blood exposures were common and cognitive outcomes were all based on the HCAP standards. Instead, we compared data qualitatively to focus on direction of effect estimates to triangulate conclusions. Results between cohorts were in agreement and we describe evidence of common markers that are not related to cultural differences.

The LASI-DAD cohort is designed to represent a broad section of the Indian population. Respondents from rural communities and representation across states means inferences made here are based on a cohort collected from wide ranging geographic regions. Additionally, the use of detailed cognitive assessment allows us to understand patient capacity and severity of condition. However, this study describes a cross-sectional analysis of the LASI baseline data and therefore we cannot determine causality. However, as the LASI-DAD cohort expands to include new timepoints it will be possible to run longitudinal analysis to examine the causal relationship between anemia with cognitive decline and dementia risk.

Recent studies of anemia in the Indian women of reproductive age have questioned the suitability of the global WHO clinical definition and suggest these guidelines may inflate prevalence in the female population ^25^. Accordingly, the study reports analysis on hemoglobin concentration and other blood indices to support any results using the clinical anaemia phenotype. Here, we describe two distinct cohorts with common cognitive test domains. We used general cognitive score for comparing to blood count measures and across cohorts as this had a high marginal reliability between cohorts when harmonised ^23^.

## Conclusion

We have established for the first time in a large South Asian population that anemia may be an important potentially modifiable risk factor for low cognitive function in an elderly population. Detection of anemia and low hemoglobin concentration is possible using common blood count assays making clinical assessment for the condition accessible and broadening research possibilities and impact particularly for dementia prevention. Increased prevalence anemia in South Asian cohorts makes further work into the effects and causes an important next step.

## Data Availability

All data produced in the present work are contained in the manuscript.

## Abbreviations

BMI: Body mass index
HCAP: Harmonized Cognitive Assessment Protocol
HRS: Health and Retirement Study
LASI-DAD: Longitudinal Aging Study in India - Harmonized Diagnostic Assessment of Dementia
MCV: Mean corpuscular volume
RDW: Red cell distribution width
RBC: Red blood cell
MCHC: Mean corpuscular hemoglobin concentration
MCH: Mean corpuscular hemoglobin
HGB: Hemoglobin concentration
HCT: Hemocrit
WHO: World Health Organisation

## Acknowledgements

We would like to thank all LASI and LASI-DAD team members and collaborators for their contribution to the wave 1 data collection.

## Funding

LW is funded by Alzheimer’s Research UK (ARUK-RF2020A-005), Michael J Fox Foundation and Rosetrees Trust (M937). The LASI cohort and data collection is funded by the National Institute on Aging, National Institutes of Health, the USA (R01 AG042778, 2R01 AG051125 and U01AG065958), and the Ministry of Health and Family Welfare, Government of India (T22011/02/2015-NCD).

## Notes

### Competing Interest Statement

The authors have declared no competing interest.

### Funding Statement

LW is funded by Alzheimers Research UK (ARUK-RF2020A-005); Michael J Fox Foundation and Rosetrees Trust (M937). The LASI cohort and data collection is funded by the National Institute on Aging; National Institutes of Health; the USA (R01 AG042778; 2R01 AG051125 and U01AG065958); and the Ministry of Health and Family Welfare; Government of India (T22011/02/2015-NCD).

### Author Declarations

All data in the present work are available from online portals LASI-DAD: GATEWAY TO GLOBAL AGING DATA HRS website: https://hrs.isr.umich.edu/data-products

## References

1. De Benoist, B., World Health Organization, & Centers for Disease Control and Prevention (U.S.). Worldwide prevalence of anaemia 1993-2005 of: WHO Global Database of anaemia. (World Health Organization, 2008).

2. Wolters, F. J. et al. Hemoglobin and anemia in relation to dementia risk and accompanying changes on brain MRI. Neurology 10.1212/WNL.0000000000008003 (2019) doi:10.1212/WNL.0000000000008003.

3. GBD 2021 Anaemia Collaborators. Prevalence, years lived with disability, and trends in anaemia burden by severity and cause, 1990–2021: findings from the Global Burden of Disease Study 2021. Lancet Haematol. 0, (2023).

4. Pasricha, S.-R. et al. Determinants of Anemia Among Young Children in Rural India. Pediatrics 126, e140–e149 (2010).

5. Mukadam, N., Sommerlad, A., Huntley, J. & Livingston, G. Population attributable fractions for risk factors for dementia in low-income and middle-income countries: an analysis using cross-sectional survey data. Lancet Glob. Health 7, e596–e603 (2019).

6. Perianayagam, A. et al. Cohort Profile: The Longitudinal Ageing Study in India (LASI). Int. J. Epidemiol. dyab266 (2022) doi:10.1093/ije/dyab266.

7. Bloom, D. E., Sekher, T. V. & Lee, J. Longitudinal Aging Study in India (LASI): new data resources for addressing aging in India. *Nat*. Aging 1, 1070–1072 (2021).

8. Lee, J. et al. Deep phenotyping and genomic data from a nationally representative study on dementia in India. Sci. Data 10, 45 (2023).

9. Hu, P. et al. Cognitive Function and Cardiometabolic-Inflammatory Risk Factors Among Older Indians and Americans. J. Am. Geriatr. Soc. 68, (2020).

10. WHO. Haemoglobin concentrations for the diagnosis of anaemia and assessment of severity. Vitamin and Mineral Nutrition Information System. https://apps.who.int/iris/bitstream/handle/10665/85839/WHO_NMH_NHD_MNM_11.1_eng.pdf?sequence=22&isAllowed=y (2011).

11. Northrop-Clewes, C. A. & Thurnham, D. I. Biomarkers for the differentiation of anemia and their clinical usefulness. J. Blood Med. 4, 11–22 (2013).

12. Lee, J., Banerjee, J., Khobragade, P. Y., Angrisani, M. & Dey, A. B. LASI-DAD study: a protocol for a prospective cohort study of late-life cognition and dementia in India. BMJ Open 9, e030300 (2019).

13. Gross, A. L., Khobragade, P. Y., Meijer, E. & Saxton, J. A. Measurement and Structure of Cognition in the Longitudinal Aging Study in India–Diagnostic Assessment of Dementia. J. Am. Geriatr. Soc. 68, S11–S19 (2020).

14. Sonnega, A. et al. Cohort Profile: the Health and Retirement Study (HRS). Int. J. Epidemiol. 43, 576–585 (2014).

15. Fisher, P. G. G., Hassan, H., Faul, J. D., Rodgers, W. L. & Weir, D. R. Health and Retirement Study Imputation of Cognitive Functioning Measures: 1992 – 2014 (Final Release Version) Data Description.

16. Didzun, O. et al. Anaemia among men in India: a nationally representative cross-sectional study. Lancet Glob. Health 7, e1685–e1694 (2019).

17. Lee, J. et al. Education, gender, and state-level disparities in the health of older Indians: Evidence from biomarker data. Econ. Hum. Biol. 19, 145–156 (2015).

18. Jiang, Z. et al. Red Cell Distribution Width and Dementia Among Rural-Dwelling Older Adults: The MIND-China Study. J. Alzheimers Dis. 83, 1187–1198 (2021).

19. Pilling, L. C. et al. Red blood cell distribution width: Genetic evidence for aging pathways in 116,666 volunteers. PLOS ONE 12, e0185083 (2017).

20. Dong, Y. et al. Neutrophil hyperactivation correlates with Alzheimer’s disease progression. Ann. Neurol. 83, 387–405 (2018).

21. Andro, M., Le Squere, P., Estivin, S. & Gentric, A. Anaemia and cognitive performances in the elderly: a systematic review. Eur. J. Neurol. 20, 1234–1240 (2013).

22. Petersen, R. C. Mild Cognitive Impairment. Contin. Lifelong Learn. Neurol. 22, 404 (2016).

23. Gross, A. L. et al. Harmonisation of later-life cognitive function across national contexts: results from the Harmonized Cognitive Assessment Protocols. Lancet Healthy Longev. 4, e573–e583 (2023).

24. Winchester, L. M., Powell, J., Lovestone, S. & Nevado-Holgado, A. J. Red blood cell indices and anaemia as causative factors for cognitive function deficits and for Alzheimer’s disease. Genome Med. 10, 51 (2018).

25. Ghosh, S. et al. Haemoglobin diagnostic cut-offs for anaemia in Indian women of reproductive age. Eur. J. Clin. Nutr. 77, 966–971 (2023).

